## Appendix for "Integrating remote monitoring into heart failure patients’ care regimen: A pilot study"

**S1 Table. Shortened Version of MDPQ.**

| Section Title, Question No. | Question |
|---|---|
| Mobile Device Basics | |
| 1 | Using a mobile device, I can navigate onscreen menus using the touchscreen |
| 2 | Using a mobile device, I can use the onscreen keyboard to type |
| Internet | |
| 1 | Using a mobile device, I can find information about my hobbies and interests on the Internet |
| 2 | Using a mobile device, I can find health information on the Internet |
| Entertainment | |
| 1 | Using a mobile device, I can use the device's online "store" to find games and other forms of entertainment (e.g., using Apple App Store or Google Play Store) |
| 2 | Using a mobile device, I can listen to music |
| Troubleshooting and Software Management | |
| 1 | Using a mobile device, I can update games and other applications |
| 2 | Using a mobile device, I can delete games and other applications |

**S2 Table. Demographics Questionnaire in the Baseline Survey.**

| Section Title, Question No. | Question |
|---|---|
| Demographics | |
| 1 | Age |
| 2 | Sex |
| 3 | Hispanic or Spanish Origin |
| 4 | Race or Ethnicity |
| 5 | Education |
| 6 | Annual Income |

**S3 Table. Questions in the Follow-Up Surveys.**

| Section Title, Question No. | Question |
|---|---|
| Hospital Readmission | |
| 1 | Have you been hospitalized within the last 4 weeks/3 months/6 months? (Do not include Emergency Room visits where you were not admitted to the hospital.) |

| | | |
|---|---|---|
| | 2 | When was the first day of your first hospitalization? |
| | 3 | What is the name of the hospital where you were first hospitalized |
| | 4 | How many times were you hospitalized in the last 4 weeks/3 months/6 months? |
| | 5 | How many nights were you in the hospital? |
| ER | | |
| | 1 | Have you visited the emergency room within the last 4 weeks/3 months/6 months (whether or not you were admitted to the hospital)? |
| | 2 | When was the first visit to the emergency room? |
| | 3 | What is the name of the emergency room you first visited? |
| | 4 | How many visits to any emergency did you have in the last 4 weeks/3 months/6 months? |
| Device | | |
| | 1 | On a scale of 1-5, with 1 being not at all helpful and 5 being extremely helpful did you find the Fitbit? |
| | 2 | What did you like and/or dislike about the Fitbit? Follow up: how easy was incorporating the Fitbit into your daily life? |
| | 3 | Did the Fitbit help you adhere to your care plan? Why or why not? |
| | 4 | On a scale of 1-5, with 1 being not at all helpful and 5 being extremely helpful did you find the scale? |
| | 5 | What did you like and/or dislike about the scale? Follow up: how easy was incorporating the scale into your daily life? |
| | 6 | Did the scale help you adhere to your care plan? Why or why not? |
| | 7 | On a scale of 1-5, with 1 being not at all helpful and 5 being extremely helpful did you find the pill bottle cap? |
| | 8 | What did you like and/or dislike about the pill bottle cap? Follow up: how easy was incorporating the pill bottle cap into your daily life? |
| | 9 | Did the pill bottle cap help you adhere to your care plan? Why or why not? |
| Extra Question Upon Completion | | |
| | 1 | Are you willing to allow our study team to continue to access data from your monitoring devices? |

### S4 Table: Questionnaires Scores at Baseline and Follow-Up.

| Questionnaire | Baseline (*n*) | Median Score (IQR) | Follow-up (n) | Median Score (IQR) |
|---|---|---|---|---|
| **Self-Care of Heart Failure Index** | | | | |
| Maintenance | 13 | 70.0 (52.6-75.0) | 14 | 76.7 (70.0-93.3) |
| Management | 12 | 62.5 (45.0-75.0) | 8 | 62.5 (47.5-72.5) |
| Confidence | 13 | 72.3 (58.4-91.8) | 14 | 58.4 (44.5-66.7) |
| Seattle Angina Questionnaire | 12 | 56.4 (51.1-69.2) | 6 | 62.8 (55.3-79.3) |
| Kansas City Cardiomyopathy Questionnaire | 13 | 45.7 (39.3-60.7) | 14 | 67.9 (47.1-85.7) |
| **PROMIS Global Health** | | | | |
| Physical | 12 | 41.1 (34.9-49.3) | 14 | 39.8 (37.4-42.3) |

| | | | | |
|---|---|---|---|---|
| Mental | 12 | 45.9 (42.3-57.5) | 14 | 45.8 (43.5-48.3) |
| PROMIS Physical Function | 13 | 35.4 (31.3-40.0) | 14 | 40.1 (31.0-45.2) |
| PROMIS Fatigue | 13 | 62.7 (57.9-64.6) | 14 | 52.1 (48.6-62.7) |
| PROMIS Anxiety | 13 | 54.2 (48.8-60.7) | 14 | 48.8 (39.1-52.7) |
| PROMIS Depression | 12 | 51.8 (41.0-57.3) | 14 | 49.0 (41.0-55.7) |
| PROMIS Sleep Disturbance | 12 | 57.0 (54.3-60.8) | 14 | 57.0 (50.5-59.8) |
| PROMIS Social Isolation | 12 | 44.5 (34.8-50.8) | 14 | 43.3 (34.8-47.8) |

### S5 Table. Ratings of the Study Devices.

| Question | *n* | Median Score (IQR) |
|---|---|---|
| On a scale of 1-5, with 1 being not at all helpful and 5 being extremely helpful, how helpful did you find the Fitbit? | 13 | 4.0 (3.0-5.0) |
| On a scale of 1-5, with 1 being not at all helpful and 5 being extremely helpful, how helpful did you find the scale? | 14 | 4.0 (4.0-5.0) |
| On a scale of 1-5, with 1 being not at all helpful and 5 being extremely helpful, how helpful did you find the pill bottle cap? | 14 | 2.0 (1.0-3.0) |

### S6 Table. Usage of the Study Devices at 30, 90, and 180 Days After Discharge.

| | 30-day | 90-day | 180-day |
|---|---|---|---|
| HR-hour (*n* = 10) | 86.0% (54.7%-95.1%) | 78.8% (64.8%-91.6%) | 80.6% (52.7%-85.6%) |
| HR-minute (*n* = 10) | 83.9% (48.5%-92.7%) | 75.3% (60.9%-90.6%) | 76.5% (49.4%-81.6%) |
| Scale (*n* = 10) | 86.7% (73.3%-100.0%) | 78.9% (60.0%-91.1%) | 66.4% (45.6%-84.3%) |
| Pill Bottle (*n* = 10) | 11.7% (0.0%-46.7%) | 3.9% (0.0%-20.0%) | 2.0% (0.0%-30.0%) |

### S7 Table. Questionnaires Scores at Baseline and 30-Day Follow-Up.

| Questionnaire | Baseline (*n*) | Median Score (IQR) | 30-day (*n*) | Median Score (IQR) |
|---|---|---|---|---|
| **Self-Care of Heart Failure Index** | | | | |
| Maintenance | 9 | 63.3 (45.0-75.0) | 10 | 76.7 (70.0-86.7) |
| Management | 8 | 50.0 (35.0-70.0) | 5 | 60.0 (42.5-65.0) |
| Confidence | 9 | 72.3 (52.8-83.4) | 10 | 52.8 (38.9-61.2) |
| Seattle Angina Questionnaire | 9 | 53.2 (44.7-73.4) | 5 | 68.1 (56.4-85.1) |
| Kansas City Cardiomyopathy Questionnaire | 10 | 48.6 (34.3-64.3) | 10 | 54.3 (47.1-75.7) |
| **PROMIS Global Health** | | | | |
| Physical | 9 | 39.8 (33.7-46.6) | 10 | 38.6 (34.9-47.7) |
| Mental | 9 | 43.5 (40.0-53.7) | 9 | 45.8 (41.2-48.3) |
| PROMIS Physical Function | 10 | 33.6 (30.3-40.0) | 10 | 34.5 (31.0-40.0) |
| PROMIS Fatigue | 10 | 62.7 (55.1-64.6) | 10 | 58.9 (55.1-64.6) |
| PROMIS Anxiety | 10 | 49.9 (48.8-55.6) | 10 | 39.1 (39.1-50.9) |
| PROMIS Depression | 9 | 53.9 (41.0-57.3) | 10 | 51.8 (41.0-57.3) |
| PROMIS Sleep Disturbance | 9 | 54.3 (52.4-58.9) | 10 | 51.5 (41.1-57.9) |
| PROMIS Social Isolation | 9 | 49.8 (34.8-51.8) | 10 | 44.5 (34.8-51.8) |

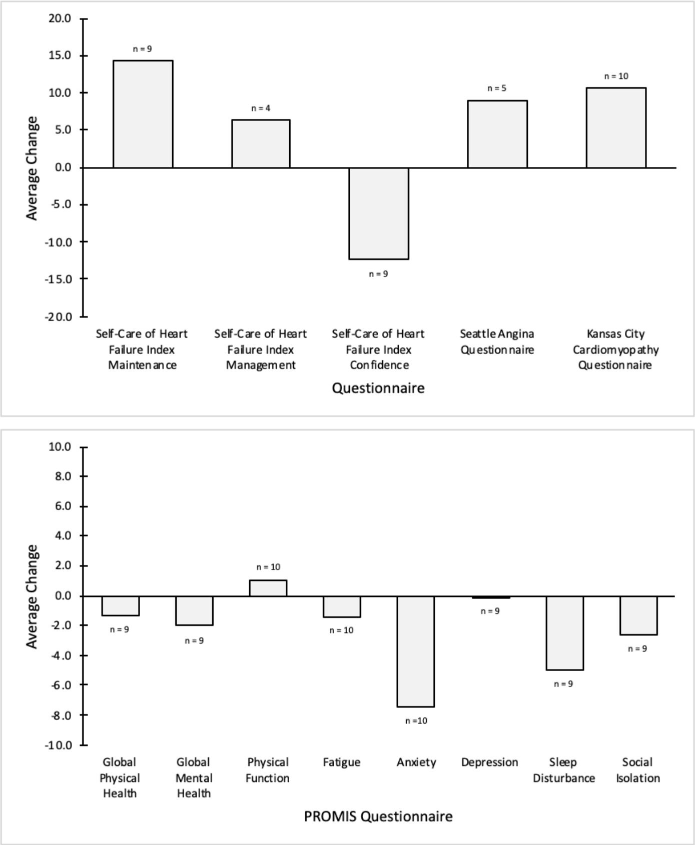


**S1 Fig. Average Changes in Patient-Reported Outcomes 30 Days After Discharge.** For non-PROMIS questionnaires, a positive change indicates improvement in health status. A positive change also signifies improvement in health status for the following PROMIS questionnaires: Global Physical Health, Global Mental Health, and Physical Function. Conversely, a negative change is indicative of improvement in health status for the following PROMIS questionnaires: Fatigue, Anxiety, Depression, Sleep Disturbance, and Social Isolation.

**S8 Table. Questionnaires Scores at Baseline, 90-Day Follow-Up, and 180-Day Follow-Up.**

| Questionnaire | Baseline (*n*) | Median Score (IQR) | 90-day (n) | Median Score (IQR) | 180-day (n) | Median Score (IQR) |
|---|---|---|---|---|---|---|
| **Self-Care of Heart Failure Index** | | | | | | |
| Maintenance | 6 | 66.7 (53.3-76.7) | 7 | 76.7 (60.0-90.0) | 7 | 73.3 (70.0-90.0) |
| Management | 6 | 70.0 (60.0-75.0) | 2 | 62.5 (45.0-80.0) | 3 | 50.0 (45.0-80.0) |
| Confidence | 6 | 80.7 (50.0-100.0) | 7 | 44.5 (44.5-66.7) | 7 | 61.2 (44.5-66.7) |
| Seattle Angina Questionnaire | 5 | 55.3 (51.1-74.5) | 5 | 91.5 (64.9-94.7) | 1 | 55.3 (NA) |
| Kansas City Cardiomyopathy Questionnaire | 6 | 45.0 (34.3-51.4) | 7 | 80.0 (48.6-84.3) | 7 | 82.9 (44.3-85.7) |
| **PROMIS Global Health** | | | | | | |
| Physical | 6 | 37.4 (32.4-42.3) | 7 | 39.8 (37.4-47.7) | 7 | 37.4 (34.9-42.3) |
| Mental | 6 | 42.3 (38.8-48.3) | 6 | 43.5 (41.1-53.3) | 7 | 43.5 (38.8-45.8) |
| PROMIS Physical Function | 6 | 32.6 (30.3-41.5) | 7 | 37.9 (29.6-46.5) | 7 | 41.5 (31.0-45.2) |
| PROMIS Fatigue | 6 | 64.6 (62.7-64.6) | 7 | 64.6 (47.3-64.6) | 7 | 51.0 (46.0-62.7) |
| PROMIS Anxiety | 6 | 54.2 (50.9-62.0) | 7 | 50.9 (48.8-58.2) | 7 | 48.8 (39.1-56.9) |

| | | | | | | |
|---|---|---|---|---|---|---|
| PROMIS Depression | 5 | 53.9 (45.0-59.7) | 7 | 53.9 (51.8-57.3) | 7 | 49.0 (41.0-60.5) |
| PROMIS Sleep Disturbance | 5 | 57.9 (47.7-66.6) | 7 | 54.3 (48.4-61.7) | 7 | 52.4 (43.8-63.8) |
| PROMIS Social Isolation | 5 | 49.8 (34.8-52.9) | 7 | 47.8 (40.3-53.9) | 7 | 43.3 (43.3-53.9) |

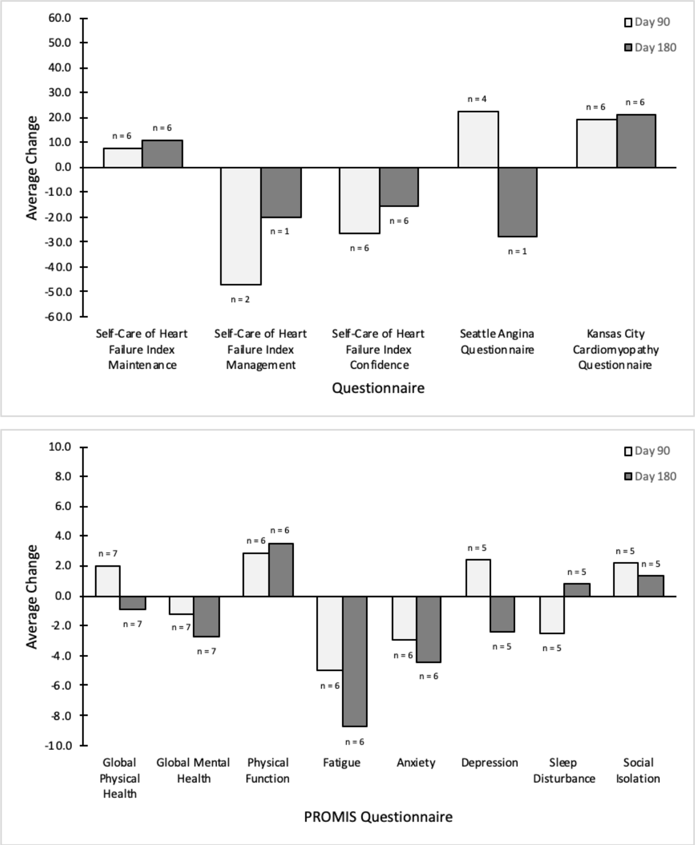


**S2 Fig. Average Changes in Patient-Reported Outcomes 90 and 180 Days After Discharge.** For non-PROMIS questionnaires, a positive change indicates improvement in health status. A positive change also signifies improvement in health status for the following PROMIS questionnaires: Global Physical Health, Global Mental Health, and Physical Function. Conversely, a negative change is indicative of improvement in health status for the following PROMIS questionnaires: Fatigue, Anxiety, Depression, Sleep Disturbance, and Social Isolation.
